# Barriers to help-seeking for Malaysian women with symptoms of breast cancer: a mixed-methods, two-step cluster analysis

**DOI:** 10.1101/2022.05.19.22275285

**Authors:** Nadia Rajaram, Maheswari Jaganathan, Kavitha Muniandy, Yamuna Rajoo, Hani Zainal, Norlia Rahim, Nurul Ain Tajudeen, Nur Hidayati Zainal, Azuddin Mohd Khairy, Mohamed Yusof Abdul Wahab, Soo Hwang Teo

## Abstract

**Objectives:** Improving help-seeking behaviour is a key component of down-staging breast cancer and improving survival, but the specific challenges faced by low-income women in an Asian setting remain poorly characterized. Here, we determined the extent of help-seeking delay among Malaysian breast cancer patients and explored sub-groups of women who may face specific barriers.

**Methods:** Time to help-seeking was assessed in 303 women diagnosed with advanced breast cancer between January 2015 and March 2020 at a suburban tertiary hospital in Malaysia. Two-step cluster analysis was conducted to identify subgroups of women who share similar characteristics and barriers. Barriers to help-seeking were identified from nurse interviews and were analyzed using behavioural frameworks.

**Results:** The average time to help-seeking was 65 days (IQR = 250 days), and up to 44.5% of women delayed by at least 3 months. Three equal-sized clusters emerged with good separation by time to help-seeking (p<0.001). The most reported barrier across clusters was poor knowledge (36.3%), regardless of help-seeking behaviour (p=0.931). Unexpectedly, women with no delay (9 days average) and great delay (259 days average) were more similar to each other than to women with mild delays (58 days average). In contrast, women who experienced great delay reported poor motivation (p=0.066) and social opportunities (p=0.374) to seek help.

**Conclusions:** Down-staging of breast cancer in Malaysia will require a multi-pronged approach aimed at alleviating culturally specific social and emotional barriers, eliminating misinformation, and instilling motivation to seek help for breast health for the women most vulnerable to help-seeking delays.

## Introduction

Although breast cancer incidence is higher in high income countries (HICs), incidence rates have been rising steadily in the majority of low-or middle-income countries (LMICs) (1). For example, in Asia, breast cancer incidence has risen by up to 6% annually between 1998 and 2012 (1). Furthermore, overall survival from breast cancer is lower in LMICs, averaging 65-85% compared to more than 90% in most HICs (2). Taken together, these statistics highlight the need to develop effective cancer control plans for resource-constrained healthcare systems (3,4).

Lower survival rates are largely attributed to late presentation of breast cancer as well as lack of timely and adequate access to treatment (5,6). The WHO Framework for Cancer Control describes the patient journey in 3 actionable intervals, which includes the presentation interval (symptom awareness to first encounter with healthcare), the diagnosis interval (first encounter to confirmed diagnosis), and the treatment interval (diagnosis to treatment onset) (7). Compared to the latter two intervals, the presentation interval may have the most influence on the stage at diagnosis, and delays in this interval could increase 5-year mortality risk by 12% (6). The presentation interval is also most likely driven by modifiable patient factors, making it an ideal target for intervention to down-stage breast cancer (8).

There is current extensive research on factors that are associated with delays in the presentation interval, or help-seeking delays, in both HICs and LMICs (8–10). Overarchingly, common reasons for delay include poor health literacy or breast cancer awareness, fear, social influences, and socioeconomic barriers. Some themes appear to be more common in LMICs, such as health care access barriers, cancer stigma, and sociocultural factors including use of alternative or traditional treatments (8,10). Notably, much of the research on help-seeking delay have investigated these determinants or barriers as acting independently (9,11), and few have used behavioural frameworks to tease out the complex and dynamic barriers that women face in seeking help for their breast symptoms (10).

To address these gaps for Malaysian women, we sought to determine the average time to help-seeking for breast symptoms among Malaysian women presenting with Stage 3 and 4 breast cancer. Using established behavioural frameworks, we sought to identify the underlying barriers to help-seeking in this population and explored for subgroups of women who may face specific barriers.

## Methodology

### Sample selection

As part of the Patient Navigation Programme (12), women with suspicious breast symptoms were referred to the Pink Ribbon Centre at a suburban tertiary hospital in Malaysia (Tengku Ampuan Rahimah Hospital, Klang) for diagnosis and treatment. A total of 371 newly diagnosed breast cancer patients presented with advanced disease (Stage 3 and 4) between January 2015 and March 2020. Women were excluded if they died prior to the interview (n=12), were not Malaysian (n=18), or if there was missing data (n=38). The final analysis included 303 patients.

Ethical approval for The Patient Navigation Programme was obtained from the Malaysian Medical Research Ethics Committee (NMRR-17-2951-35223), Ministry of Health Malaysia, and conforms to the principles of the Declaration of Helsinki. Informed consent is not available because data was collected as part of patient’s clinical care within the programme. Permission to use patient data for analysis was sought from the hospital under the governance of the Malaysian Ministry of Health.

### Data collection

Patients were interviewed by a trained nurse responsible for their care. Open-ended questions were used to capture barriers and help-seeking behaviour. Additionally, a structured questionnaire was used to collect socio-demographic status, medical history, breast cancer risk factors, and risk management behaviour. The questionnaire was developed by incorporating best practices in patient intake assessments as well as factors that are relevant to breast cancer risk and help-seeking behaviour among Malaysian women. It was tested in a pilot study of 30 breast cancer patients prior to data collection *(data not published)*.

### Statistical analysis

Standard descriptive statistics were used to describe the distribution of patients by demographic and socio-economic factors, knowledge, screening, barriers, and time to help-seeking. Time to help seeking was defined from the date of symptom awareness to the date of first medical encounter for the symptom. If the date was missing the day or month information, we assumed it to be the 1^st^ of the month or June of that year, respectively.

A multivariable logistic regression model was used to assess the association between patient factors and time to help-seeking (≤ 3 months vs > 3 months) (6). Next, two-step cluster analysis was performed on the dataset using the algorithm in the IBM® SPSS® Statistics software. Using backward elimination, variables were removed if they were poorly associated with time to help-seeking in the regression model (lenient p>0.600) or showed poor predictor importance in two-step cluster analysis (<15%, Supplementary Table 1). The model was finalized when suitable clustering parameters were achieved (Supplementary Table 1). Internal validation of the cluster analysis was conducted using principal component analysis (Supplementary Figure 1).

Differences across clusters were assessed with global Kruskal-Wallis tests for non-parametric continuous variables and independent Chi-square tests for homogeneity for categorical variables. Variables with p<0.100 in global tests were subjected to pairwise Wilcoxon tests with Benjamini-Hochberg correction or Chi-square tests with Bonferroni correction, respectively. These analyses were performed using the R Statistical environment (v4.0.3).

### Thematic analysis

Barriers to help-seeking were identified from notes collated by the nurse during interview with patients. These barriers were grouped into themes and further mapped to a combined matrix of two behavioural models, namely the Capability, Opportunity, and Motivation Model for Behaviour (COM-B) and the Theoretical Domains Framework (TDF) (13). This process was conducted independently by two reviewers and discrepancies were resolved by consensus. A summary of the barriers in each COM-B/TDF domain is described (Supplementary Table 2). Fisher’s Exact tests were used to test for differences in the distribution of COM-B/TDF domains across clusters. These analyses were performed using the R Statistical environment (v4.0.3).

All hypotheses were two-sided, and p<0.05 was considered statistically significant.

## Results

### Cohort description and time to help seeking

In this cross-sectional analysis of 303 newly diagnosed Malaysian breast cancer patients presenting with advanced disease, the average time to help-seeking was approximately 2 months (median=65 days, IQR=250 days). Almost half of women (n=136, 44.5%) delayed help-seeking by >3 months. Also, as shown in Figure 1, there was great variation in time to help-seeking in the overall cohort, with 18% of the patients reporting >1 year delay.

**Figure 1:**
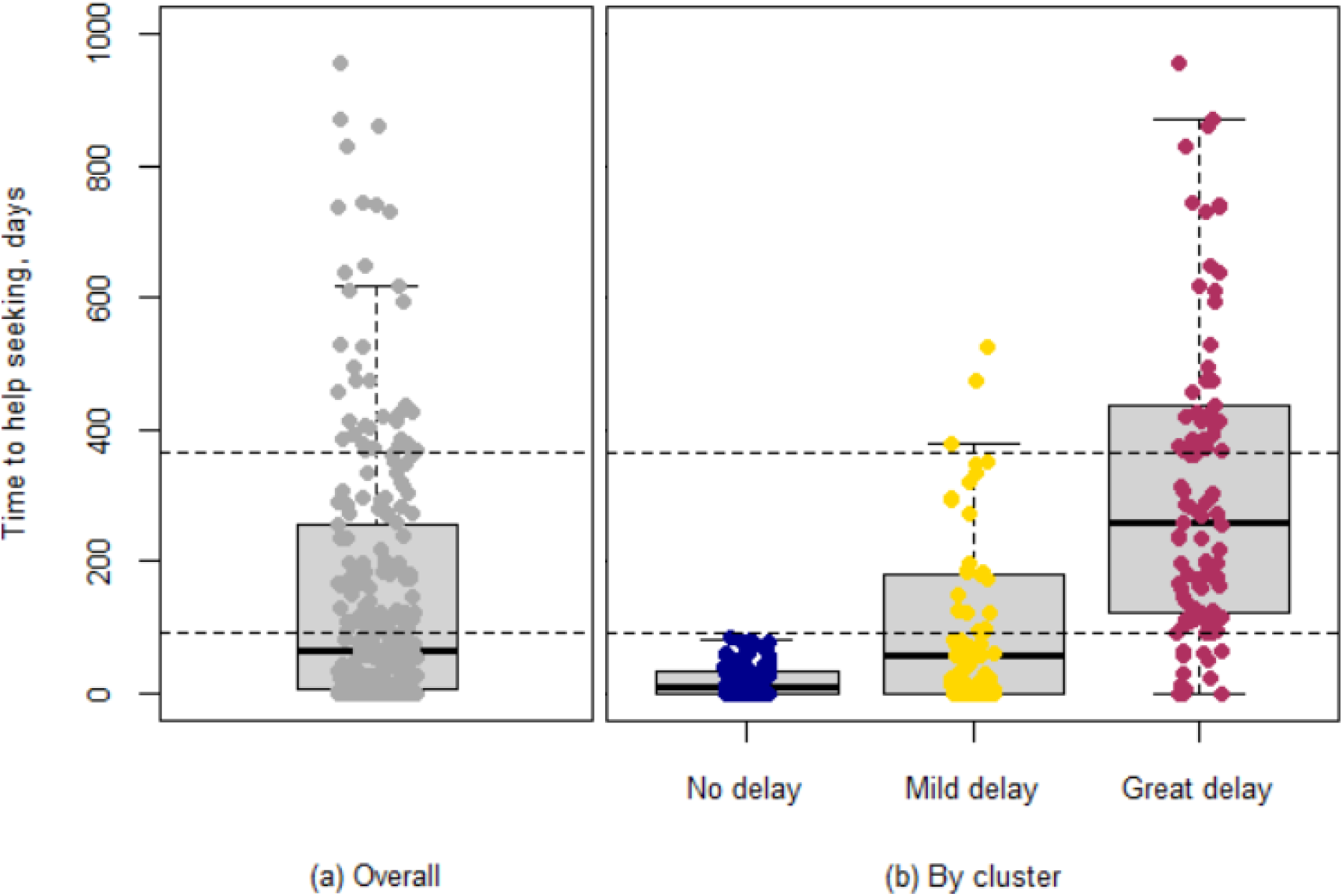
Distribution of time to help seeking (a) overall and (b) by cluster.

A low proportion of women reported knowing the signs and symptoms (42.6%) or risk factors (16.5%) of breast cancer (Table 1). Screening behaviour was also poor, with only 17.8% reporting ever being screened for breast cancer. With regards to help-seeking behaviour, most women (64.4%) reported shared decision making, whilst 13% of women reported that they make their own health decisions. For up to 21% of women, however, their health decisions are made by their family. Furthermore, nearly one-third (29%) of women reported a past bad experience with healthcare or a life crisis. Up to 43% of women lived more than 10km from the hospital and 26.7% reported transportation as a barrier to seeking healthcare services. A small minority reported a language barrier when communicating with hospital staff (6.9%).

**Table 1:** Demographics of breast cancer patients, by time to help-seeking

| Characteristics | Overall<br>(n=303) | By time to help-seeking,<br>median±IQR or n(%) |  | Multivariable analysis |  |
| --- | --- | --- | --- | --- | --- |
|  |  | ≤3 months<br>(n=167) | >3months<br>(n=136) | OR[95%CI] | p-value |
| **Demographic factors** |  |  |  |  |  |
| Age, years | 55.1±14.0 | 54.5±14.2 | 55.9±13.8 | 0.01[0.02,0.05] | 0.455 |
| Ethnicity |  |  |  |  |  |
| Malay | 179(59.1) | 90(53.9) | 89(65.4) | Ref |  |
| Indian | 77(25.4) | 50(29.9) | 27(19.9) | -0.60[-1.58,0.35] | 0.226 |
| Chinese | 47(15.5) | 27(16.2) | 20(14.7) | 0.03[-0.95,0.99] | 0.957 |
| Marital status |  |  |  |  |  |
| Married | 181(59.7) | 103(61.7) | 78(57.4) | Ref |  |
| Divorced/widowed | 86(28.4) | 41(24.6) | 45(33.1) | 0.44[-0.46,1.34] | 0.340 |
| Single | 35(11.6) | 22(13.2) | 13(9.6) | -0.36[-1.54,0.77] | 0.535 |
| Children |  |  |  |  |  |
| None | 50(16.5) | 30(18.0) | 20(14.7) | * |  |
| 1-3 | 145(47.9) | 76(45.5) | 69(50.7) |  |  |
| >3 | 107(35.5) | 61(36.5) | 46(33.8) |  |  |
| **Socio-economic factors** |  |  |  |  |  |
| Currently employed |  |  |  |  |  |
| No | 220(72.6) | 121(72.5) | 99(72.8) | Ref |  |
| Yes | 77(25.4) | 42(25.1) | 35(25.7) | 0.18[-0.69,1.04] | 0.686 |
| Household income |  |  |  |  |  |
| ≥RM800/month | 205(67.7) | 115(68.9) | 90(66.2) | Ref |  |
| <RM800/month | 89(29.4) | 49(29.3) | 40(29.4) | 0.37[-0.44,1.19] | 0.374 |
| Formal education |  |  |  |  |  |
| Incomplete | 180(59.4) | 103(61.7) | 77(56.6) | Ref |  |
| Secondary/tertiary | 120(39.6) | 61(36.5) | 59(43.4) | 0.27[-0.57,1.11] | 0.532 |
| **Knowledge & screening<sup>†</sup>** |  |  |  |  |  |
| Signs & symptoms | 129(42.6) | 61(36.5) | 68(50.0) | 0.71[-0.09,1.53] | 0.085 |
| Risk factors | 50(16.5) | 21(12.6) | 29(21.3) | 0.19[-0.76,1.14] | 0.694 |
| Ever screened | 54(17.8) | 28(16.8) | 26(19.1) | 0.32[-0.63,1.29] | 0.507 |
| **Barriers** |  |  |  |  |  |
| Distance to hospital |  |  |  |  |  |
| <10km | 172(56.8) | 99(59.3) | 73 (53.7) | Ref |  |
| ≥10km | 131(43.2) | 68(40.7) | 63 (46.3) | 0.29[-0.38,0.97] | 0.394 |
| Transportation barrier<sup>†</sup> | 81(26.7) | 45(26.9) | 36 (26.5) | -0.11[-0.91,0.68] | 0.783 |
| Language barrier<sup>†</sup> | 21(6.9) | 11(6.6) | 10 (7.4) | 0.38[-0.87,1.63] | 0.547 |
| Decision maker |  |  |  |  |  |
| Self | 41(13.5) | 23(13.8) | 18(13.2) | Ref |  |
| Shared | 195(64.4) | 106(63.5) | 89(65.4) | 0.04[-0.94,1.02] | 0.936 |
| Family | 63(20.8) | 37(22.2) | 26(19.1) | 0.20[-0.89,1.30] | 0.724 |
| Bad experience/crisis<sup>†</sup> | 87(28.7) | 41(24.6) | 46(33.8) | 0.14[-0.63,0.91] | 0.720 |

<sup>†</sup>Reference group in multivariable logistic regression = "No". \*Number of children was highly correlated with marital status (VIF > 3) and was therefore excluded from multivariable analysis.

We observed no significant differences when comparing the characteristics of women by time to help-seeking (≤3 months vs >3 months, Table 1).

### Distribution of patients by two-step cluster analysis grouping

We conducted a two-step cluster analysis to explore for groups of women who may share similar characteristics and experiences. It revealed three distinct clusters with good separation by time to help-seeking (p<0.001, Figure 1). The average time to help-seeking was 9 days (IQR=32.8) for women in the “No delay” cluster, 58 days (IQR=180 days) in the “Mild delay” cluster, and 259 days in the “Great delay” cluster (IQR=314 days). In the “Great delay” cluster, up to 90% of women experienced delays of >3 months.

Unexpectedly, we observed many similarities between women with no delay and women with great delay, while women with mild delay in help-seeking were characteristically different (Table 2). Compared to women with no delay and great delay, women with mild delay were older (67 vs 47 and 52 years old, respectively, p<0.001) and were less likely to be Malay (38.3% vs 63.8% and 75.2%, p<0.001). Women with mild delay also reported more socioeconomic barriers, such as low education, unemployment, and low household income (p<0.001, respectively). Furthermore, they were less likely to report knowing about signs and symptoms or risk factors of breast cancer (p<0.001, respectively).

**Table 2:** Distribution of breast cancer patients, by two-step cluster analysis grouping

| Characteristics | Distribution by cluster<sup>‡</sup>,<br>median±IQR or n(%) |  |  | p<sub>global</sub> | Pairwise comparison,<br>p-value |  |  |
| --- | --- | --- | --- | --- | --- | --- | --- |
|  | No delay<br>(n=94) | Mild delay<br>(n=81) | Great delay<br>(n=113) |  | No vs<br>Mild | Mild vs<br>Great | No vs<br>Great |
| **Time to help seeking** |  |  |  |  |  |  |  |
| In days<sup>†</sup> | 9±33 | 58±180 | 259±314 | **<0.001** |  |  |  |
| >3 months delay | 0(0) | 29(35.8) | 101(89.4) | **<0.001** | *** | *** | *** |
| **Demographic factors** |  |  |  |  |  |  |  |
| Age in years<sup>†</sup> | 47.5±16.8 | 67.0±17.0 | 52.0±13.0 | **<0.001** |  |  |  |
| Ethnicity<sup>†</sup> |  |  |  |  |  |  |  |
| Malay | 60(63.8) | 31(38.3) | 85(75.2) | **<0.001** | * | *** |  |
| Indian | 24(25.5) | 27(33.3) | 20(17.7) |  |  |  |  |
| Chinese | 10(10.6) | 23(28.4) | 8(7.1) |  |  |  |  |
| Marital status<sup>†</sup> |  |  |  |  |  |  |  |
| Married | 85(90.4) | 23(28.4) | 66(58.4) | **<0.001** | *** | *** |  |
| Divorced/Widowed | 2(2.1) | 47(58.0) | 32(28.3) |  | *** |  | *** |
| Single | 7(7.4) | 11(13.6) | 15(13.3) |  |  |  |  |
| No. of children |  |  |  |  |  |  |  |
| None | 14(14.9) | 15(18.5) | 19(16.8) | **0.043** |  |  |  |
| 1-3 | 51(54.3) | 27(33.3) | 58(51.3) |  |  | ** |  |
| >3 | 29(30.9) | 39(48.1) | 35(31.0) |  |  |  |  |
| **Socio-economic factors** |  |  |  |  |  |  |  |
| Currently employed | 30(31.9) | 8(9.9) | 37(32.7) | **<0.001** | ** | *** |  |
| Household income |  |  |  |  |  |  |  |
| ≥RM800/month | 92(97.9) | 8(9.9) | 102(90.3) | **<0.001** | *** | *** |  |
| <RM800/month | 2(2.1) | 73(90.1) | 11(9.7) |  | *** | *** |  |
| Formal education<sup>†</sup> |  |  |  |  |  |  |  |
| Incomplete | 47(50.0) | 75(92.6) | 48(42.5) | **<0.001** |  |  |  |
| Secondary/tertiary | 47(50.0) | 6(7.4) | 65(57.5) |  | *** | *** |  |
| **Knowledge & screening** |  |  |  |  |  |  |  |
| Know sign/symptoms<sup>†</sup> | 47(50.0) | 12(14.8) | 67(59.3) | **<0.001** | *** | *** |  |
| Know risk factors | 16(17.0) | 2(2.5) | 30(26.5) | **<0.001** | ** | *** |  |
| Ever screened | 20(21.3) | 5(6.2) | 26(23.0) | **0.006** | ** | ** |  |
| **Barriers** |  |  |  |  |  |  |  |
| Distance to hospital<sup>†</sup> |  |  |  |  |  |  |  |
| <10km | 50(53.2) | 53(65.4) | 61(54.0) | 0.190 |  |  |  |
| ≥10km | 44(46.8) | 28(34.6) | 52(46.0) |  |  |  |  |
| Transportation barrier | 24(25.5) | 29(35.8) | 25(22.1) | 0.062 |  |  |  |
| Language barrier | 3(3.2) | 9(11.1) | 6(5.3) | 0.076 |  |  |  |
| Decision maker |  |  |  |  |  |  |  |
| Self | 13(13.8) | 11(13.6) | 15(13.3) | 0.340 |  |  |  |
| Shared | 64(68.1) | 46(56.8) | 76(67.3) |  |  |  |  |
| Family | 17(18.1) | 23(28.4) | 19(16.8) |  |  |  |  |
| Bad experience/crisis<sup>†</sup> | 12(12.8) | 31(38.3) | 43(38.1) | **<0.001** | * |  | *** |

<sup>†</sup>Variables included in the two-step cluster analysis model. <sup>‡</sup>A total of 15 women (5%) were not clustered due to missing data. Pair-wise p-values: \*\*\* p-value > 0.001, \*\* p-value > 0.05, \*p-value > 0.1.

Interestingly, we found that marital status and past bad experiences may be associated with delayed help-seeking behaviour in this cohort (Table 2). Compared to women with no delay, women with mild and great delay were more likely to be divorced or widowed (58.0% and 28.3% vs 2.1% among no delays, p<0.001). Up to 38% of women in the two delay clusters reported a past bad experience in healthcare or a life crisis, compared to only 12.8% among women with no delays (p<0.001).

### Barriers to help seeking among Malaysian women

Using theoretical frameworks, we mapped the barriers to help-seeking for Malaysian women presenting with advanced breast cancer into a matrix of 3 COM-B domains and 10 TDF domains (Table 3). Lack of knowledge, in the capability domain, was the most common barrier to help-seeking (36.3%). Next, barriers were commonly reported in the motivation domain, such as emotional barriers (22.1%) and optimism (20.8%). Approximately 11% of patients report lack of opportunity to seek help, due to physical and social barriers.

**Table 3:**
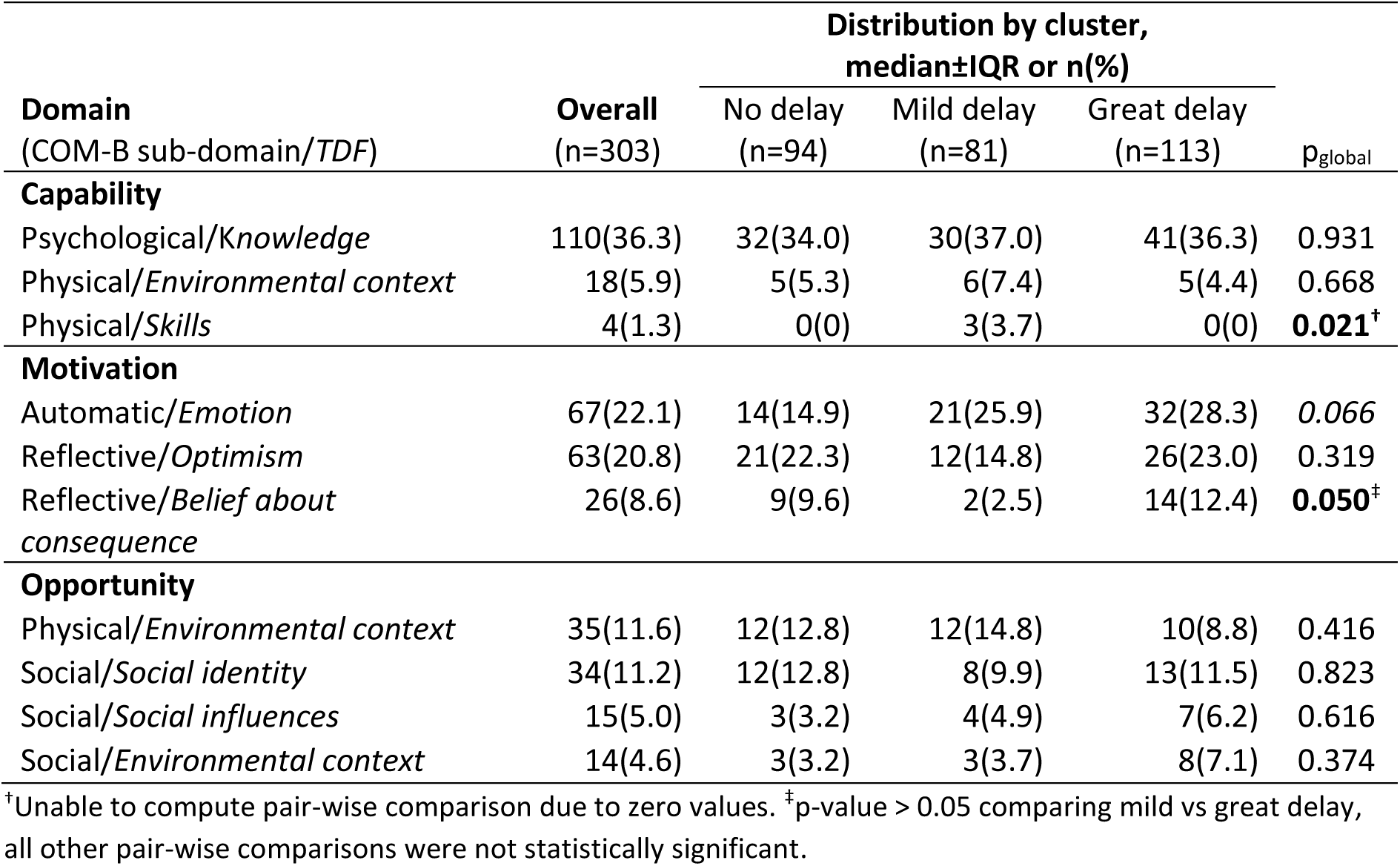
Distribution of barriers, mapped to the COM-B and TDF domains, overall and by cluster

We compared the distribution of barriers by time to help-seeking (Supplementary Table 3). Compared to women who sought help within 3 months, women who delayed help-seeking were more likely to report lack of motivation, including emotional barriers (25.7% vs 19.2%, p=0.239), optimism (24.3% vs 18.0%, p=0.251), and belief in consequences (10.3% vs 7.2%, p=0.470). Social influences were also more commonly reported among women who delayed help-seeking (7.4% vs 3.0%, p=0.147). However, these associations were not statistically significant.

When comparing the distribution of barriers by clusters, on the other hand, some similarities were observed among women with no delay and great delays (Table 3). Between 9-12% of women in these clusters reported belief in consequences as a barrier to help-seeking, compared to 2.5% among women with mild delay (p=0.050). Similarly, women with no or great delay were more likely to report optimism as a reason for delayed help-seeking but this was not significantly different from women with mild delay (22-23% vs 14.8%, p=0.319). The results observed here for women with no delay contrasts the analysis by time to help-seeking in Supplementary Table 3.

Instead, emotional and social barriers appear to be important facilitators for delayed help-seeking behaviour in this cohort. More than 25% of women with mild and great delay reported emotional barriers, compared to only 15% of women with no delay (p=0.066). Furthermore, 7% of women with the great delay reported barriers in their social environment, compared to 3-4% among women with no or mild delay, but this difference was small and not statistically significant (p=0.374). Notably, women with physical capability barriers were all clustered within the mild delay group (3.7%).

## Discussion

In this study of 303 Malaysian women presenting with advanced breast cancer, the average time from symptom discovery to help-seeking was approximately 2 months, similar previous reports in LMICs (10). Here, up to 45% of women experienced help-seeking delays by more than 3 months, and 18% were delayed by more than 1 year. Poor knowledge about breast cancer was commonly and consistently reported in this cohort of women, regardless of help-seeking behaviour. We found that women could be clustered into 3 groups based on their help-seeking behaviour. Intriguingly, instead of a graduated effect, we found that women with least and greatest delay were more similar to each other compared to women with mild delay. Importantly, we found that emotional and social barriers most likely facilitated help-seeking delay in this cohort. This study highlights that Malaysian women who face delays in seeking help are not a homogeneous group, and that varied solutions are required to effectively improve early detection of breast cancer in Malaysia.

Our study shows that poor cancer awareness remains an important barrier for Malaysian women. This is consistent with other studies that have highlighted knowledge as a primary barrier to help-seeking behaviour in both HICs and LMICs, including in Malaysia and Singapore, where women who were more aware about breast health were more likely to seek help early (9,10,14,15). As a result, the public health response to increasing early detection of breast cancer has centred on campaigns that create and raise awareness. However, the long-term efficacy of such campaigns in reducing presentation delay, and ultimately in down-staging breast cancer, is not well-studied (16). Notably, our results suggest that addressing poor knowledge alone is likely insufficient to improve early detection of breast cancer.

Instead, addressing emotional barriers may empower women to seek help early for their breast symptoms. Previous reports suggest that emotional messaging in information about breast health awareness is an important driver for help-seeking behaviour (17). Fear is a commonly used emotion in public health campaigns, including for breast cancer awareness, but it’s effect on help-seeking behaviour is inconsistent across populations (18). While some studies show that fear has a positive influence (18), we show that fear and embarrassment were important deterrents to help-seeking. This is consistent with other studies of Asian women, where fear is often reported as one of the main reasons for late presentation (15,19). Alternatively, emotional messaging with positive reinforcements that reduce the fear of treatment and death may serve as stronger motivation for Asian women and is an area of pressing unmet need that requires further investigation.

Additionally, we show that a poor social environment may preclude women of the opportunity to seek help. This finding is comparable to past studies of Asian women, where poor social support and negative social influences are often reported as primary barriers to seeking help for breast cancer symptoms (15,20,21), and may be a more significant barrier than lack of knowledge in this region (22). In Asian communities, family members appear to have both direct and indirect roles in a women’s decision to seek help (14,23,24). A supportive family can be an enabler of early help-seeking behavior and adherence to treatment (14), whereas lack of support or unstable family dynamics often lead to confusion, uncertainty, and ultimately, delay in seeking medical treatment (23). Therefore, interventions to improve screening and early detection that consider family dynamics and socio-cultural factors may prove to be more effective than a one-size-fits-all campaign (25).

Surprisingly, in our cohort, women with no delay observed similar characteristics and barriers to those with great delay. For example, we observed that women with no delay and great delay similarly reported the use of alternative treatment prior to seeking medical attention. The preference for alternative treatment is commonly reported in this region (14,20,22) as well as other LMICs (8), and has been previously shown to deter help-seeking behaviour (23). Here, we suggest that use of alternative treatment may not always lead to delays in help-seeking. Instead, delay occurs when there are other emotional and social pressures that reinforces a women’s belief that medical care is not necessary, not accessible, or not sufficient to treat her symptoms (15). This analysis further illustrates the diversity of women and the challenges that they face in seeking help for breast symptoms, which may not be elucidated in studies where barriers are assumed to act independently.

### Clinical implications

To our knowledge, this is the first study to use a mixed-method cluster analysis to understand barriers to help-seeking behavior for breast cancer symptoms. It considers the possibility that women delay help-seeking by different degrees and for various and multiple reasons (9,15). Coupling the two-step cluster analysis with thematic analysis has enabled us to study the intersections between patient characteristics and the barriers faced by different groups of women in the population. Specifically, it has identified emotional coping as the main difference between women who present with no or mild delay compared to women who have great delay, raising the intriguing possibility that public campaigns that focus on positive messaging about survival of cancer and emotional coping could have a more positive impact on help-seeking behavior than those which focus on knowledge of signs and symptoms alone.

### Study limitations

This study is not without limitations. Firstly, we have only examined patient factors and have not considered healthcare factors, such as access and coordination of care (10). Secondly, the women studied here are advanced breast cancer patients diagnosed at a suburban tertiary hospital in Malaysia. Therefore, the findings may not be inferred to all Malaysian breast cancer patients. Furthermore, patients who presented early in our study may represent a unique group of women with more aggressive disease and may not be comparable to women who present with early-stage breast cancer. Thirdly, the data used for the thematic analysis represent patient barriers identified by nurses during interviews with patients, rather than patient-reported barriers. Future research could explore whether there are differences between nurse-perceived and patient-reported barriers.

### Conclusions

Whilst poor breast cancer knowledge and awareness was commonly reported among Malaysian women, poor motivation and/or social opportunities to seek help were the primary reasons of late presentation. Understanding the unique barriers faced by specific groups of women is the first step to developing tailored interventions that can effectively reduce help-seeking delays and down-stage breast cancer in Malaysia.

## Data Availability

The data that support the findings of this study are available on request from the corresponding author. The data are not publicly available due to privacy or ethical restrictions.

## Authorship

ST, MY, AK, NR, and MJ conceptualized and designed the study. MJ, KM, HZ, NR, NT, NZ were involved in data collection and patient management at the hospital. ST, MY, NR, MJ, KV, YM conducted data analysis, interpreted the findings, and drafted the manuscript. All authors critically revised the manuscript, approved the final version for publication, and agree to be accountable for all aspect of the work published.

## Acknowledgement

We would like to thank the Director General of Health Malaysia for his permission to publish this article. We would also like to thank all parties from the Ministry of Health Malaysia who were involved in data collection and the management of patients. Lastly, we are grateful for the contributions of breast cancer patients to this study.

