## Supplementary for "Barriers to help-seeking for Malaysian women with symptoms of breast cancer: a mixed-methods, two-step cluster analysis"

Supplementary Table 1: Model selection for two-step cluster analysis

| Backward selection of variables | No. of variables | No. of clusters | Cluster quality (Silhouette index) | Cluster size ratio | Dominant variable | Separation by time to help-seeking | Suitable clustering parameters[‡] |
|---|---|---|---|---|---|---|---|
| All variables from multivariable model | 16 | 2 | Fair (0.2) | 1.27 | Age | None | No |
| Removed variables w/ p>0.600 in multivariable model | 11 | 3 | Fair (0.2) | 1.84 | None | Poor | No |
| Removed variables not important in clustering (<15% importance) [†] | 9 | 3 | Fair (0.2) | 1.40 | None | Good | Yes |

[†]Final model, variables included are age, ethnicity, marital status, income, education, knowledge of signs/symptoms, bad experience/crisis, distance to hospital, and time to help-seeking. [‡]Suitable clustering parameters are defined as at least fair cluster quality (silhouette score for cohesion and separation > 0.2), equal cluster sizes (ratio of cluster size < 3), and the absence of a dominant variable.

Supplementary Table 2: Description of themes according to COM-B and TDF domains

| COM-B domain | COM-B sub-domain | TDF domain | Description of the reasons women delay help seeking within each domain | Example quotes by nurse |
|---|---|---|---|---|
| Capability | Psychological capability | Knowledge | Don't know about breast health or symptoms of breast cancer, nor the urgency to seek help for the symptom. Also, sought help for symptom but was told it was harmless | "Patient don't know sign and symptom of breast cancer…" (Quote 1)<br>"…thought lump was [because of] breast engorgement due to lactation…" (Quote 2)<br>"Clinic general practitioner said the lump is harmless…" (Quote 3) |
| | Physical capability | Environmental context | Pregnancy or pre-existing comorbidities/ disabilities. May be physically dependent on caregivers | "Patient had long hospitalization due to pregnancy complication, …" (Quote 5)<br>"Patient with [disabilities] and depend[s] on caregiver…" (Quote 6) |
| | | Skills | Unable to read or has a language barrier, or unable raise concerns about her own health | "…illiterate, unable to communicate effectively with health care professionals…" (Quote 4) |
| Motivation | Automatic motivation | Emotion | Fear of diagnosis, treatment, and/or healthcare professionals. Fears death is inevitable. Embarrassment to discuss the symptoms with her family/peers. | "Patient is scared to go to hospital…" (Quote 7)<br>"Very shy and scared to inform family" (Quote 8)<br>"…think being punished by god… fear…" (Quote 9) |
| | Reflective motivation | Optimism | Assumed that the symptom was not harmful and can be managed at home or by alternative/traditional treatment | "…lump was painless, [patient] was self-monitoring…" (Quote 10)<br>"Started with alternative treatment…" (Quote 11) |
| | | Belief about consequences | In denial or doesn't believe that she is at risk for breast cancer. Also, may understand that she may be at risk, but is not ready or does not want to seek help | "…don't feel that she may get breast cancer as [she had] no family history of breast cancer…" (Quote 12)<br>"Patient not ready to see doctor…" (Quote 13) |

Supplementary Table 2: Description of themes according to COM-B and TDF domains *(continued)*

| COM-B domain | COM-B sub-domain | TDF domain | Description of the reasons women delay help seeking within each domain | Example quotes by nurse |
|---|---|---|---|---|
| Opportunity | Physical opportunity | Environmental context | Faces financial and/or logistical difficulties and may be dependent on others for money to seek help or transport to the hospital. | "Not working, […] depends on husband to go for follow up…" (Quote 14)<br>"…only [receives] pocket money from children, [lives far away from the hospital], and depends on other for transportation" (Quote 15) |
| | Social opportunity | Social identity | Has other priorities or commitments, i.e. work or family commitments. Worries that her symptoms or diagnosis will burden her family. | "Patient has commitment in caring for her children and husband…" (Quote 16)<br>"Doesn't want to trouble [her] children…" (Quote 17) |
| | | Social influences | In a dilemma or in confusion over conflicting, negative information from her social circle. Family is hesitant to seek help, advices her to manage at home and is supportive of alternative/traditional medicine. | "Husband [did] not allow, initiated alternative treatment first…" (Quote 18)<br>"Husband [did] not allow, [patient is in a] confused state of mind" (Quote 19)<br>"Daughter wants to monitor at home…" (Quote 20) |
| | | Environmental context | Lives alone and may be single or recently divorced. Also, has no support from family or has concomitant family problems | "Husband controlling her and [there is] no support…" (Quote 21)<br>"High family commitment, [but] no family support" (Quote 22) |

Supplementary Table 3: Distribution of barriers, mapped to the COM-B and TDF domains, by time to help seeking

| **Domain** (COM-B sub-domain/*TDF*) | **Overall** (n=303) | By time to help-seeking, n(%) ≤3 months (n=167) | >3 months (n=136) | p-value |
|---|---|---|---|---|
| **Capability** | | | | |
| Psychological/*Knowledge* | 110(36.3) | 60(35.9) | 50(36.8) | 1.000 |
| Physical/*Environmental context* | 18(5.9) | 10(6.0) | 8(5.9) | 1.000 |
| Physical/*Skills* | 4(1.3) | 2(1.2) | 2(1.5) | 1.000 |
| **Motivation** | | | | |
| Automatic/*Emotion* | 67(22.1) | 32(19.2) | 35(25.7) | 0.239 |
| Reflective/*Optimism* | 63(20.8) | 30(18.0) | 33(24.3) | 0.251 |
| Reflective/*Belief about consequence* | 26(8.6) | 12(7.2) | 14(10.3) | 0.470 |
| **Opportunity** | | | | |
| Physical/*Environmental context* | 35(11.6) | 21(12.6) | 14(10.3) | 0.635 |
| Social/*Social identity* | 34(11.2) | 19(11.4) | 15(11.0) | 1.000 |
| Social/*Social influences* | 15(5.0) | 5(3.0) | 10(7.4) | 0.147 |
| Social/*Environmental context* | 14(4.6) | 6(3.6) | 8(5.9) | 0.518 |

(a) PCA separation plot, labelled with two-step cluster analysis groups

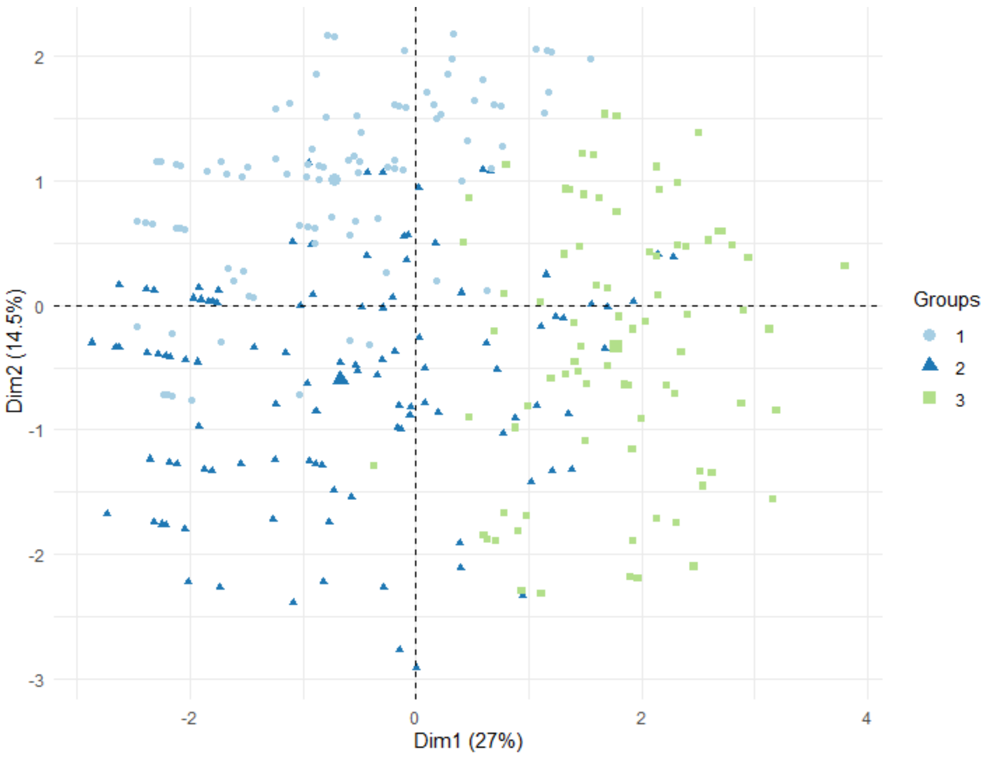


(b) Relative contribution of each variable to separation in PCA

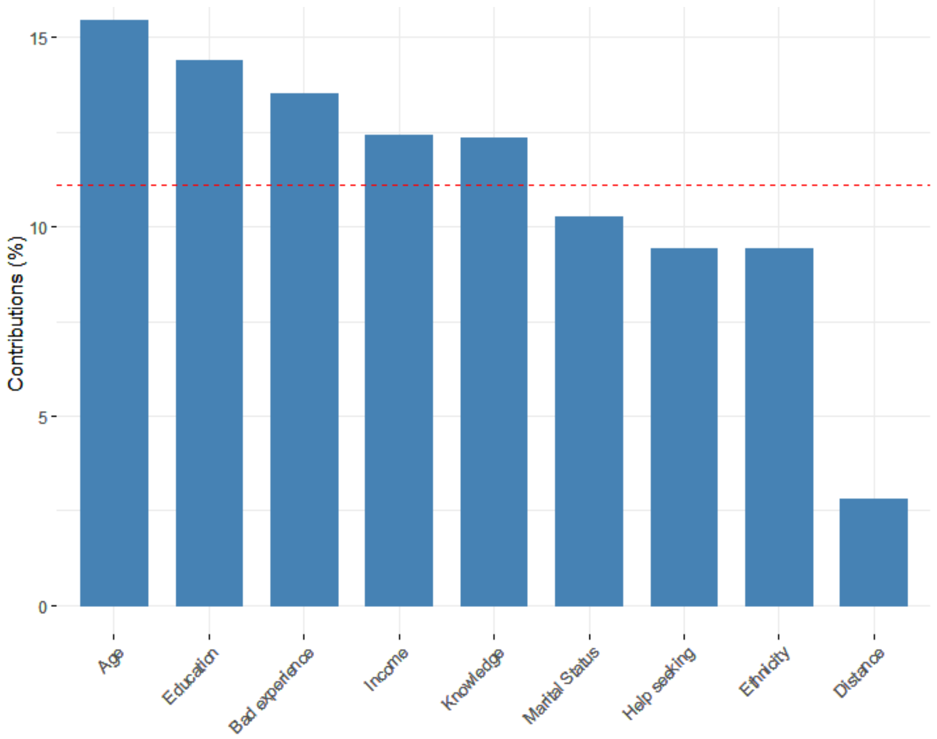


Supplementary Figure 1: Internal validation of clustering using principal component analysis (PCA) showing (a) separation plot labelled with two-step cluster analysis groups, and (b) relative contribution of each variable to separation.
